# Single-cell characterization of menstrual fluid at homeostasis and in endometriosis

**DOI:** 10.1101/2024.05.06.24306766

**Authors:** Petra C. Schwalie, Cemsel Bafligil, Julie Russeil, Magda Zachara, Marjan Biocanin, Daniel Alpern, Evelin Aasna, Bart Deplancke, Geraldine Canny, Angela Goncalves

## Abstract

Progress in detecting and understanding endometrial conditions in women of fertile age, such as endometriosis, has been hampered by the invasiveness of the sample collection procedure. Menstrual fluid (MF) can be sampled non-invasively and could provide a unique opportunity to study the physiological state of tissues in the reproductive system. Despite this potential, the use of MF for diagnostics and research has been limited. Here we establish protocols and assess the feasibility of collecting and processing MF in an outpatient setting. We characterize the cellular contents of MF from 15 healthy women using flow cytometry and single-cell RNA-sequencing, and demonstrate the ability to recover millions of live cells from the different cellular fractions of interest (epithelial, stromal, endothelial, perivascular and blood). Through computational integration of MF with endometrial samples we show that MF sampling is a good surrogate for endometrial biopsy. In a proof-of-principle case-control study, we collect MF from a further 7 women with a diagnosis of endometriosis and 11 healthy controls. Through RNA sequencing of 93 MF samples from these women we highlight important differences between *ex vivo* and cultured cells, identify impaired decidualisation, low apoptosis, high proliferation, and both higher and lower inflammatory activity in different subsets of immune cells as distinguishing features of endometriosis patients. Finally, we identify potential novel pan-cell-type biomarkers for this neglected condition.

## Introduction

Menstrual fluid (MF) is an accessible source of large numbers of live cells of high biomedical interest. During menstruation, the female body sheds 86±48ml of MF (1) containing cervico-vaginal secretions, blood and cells from the lining of uterus, cervix, vagina, and possibly the fallopian tube and ovaries. This rich source of live cells has been previously proposed as a scalable source of live uterine NK cells (2), mesenchymal stem cells (3–7), T-cells (8) and epithelial cells (9,10) for research and therapy.

Menstrual fluid has also been proposed as a sampling strategy for potential diagnosis of endometriosis (11–14), a chronic, currently incurable disease that affects an estimated 10% of women of reproductive age. Endometriosis occurs when cells similar to those that line the inside of the uterus – the eutopic endometrium – grow outside the uterine cavity in so called ectopic lesions. These ectopic, endometrial-like cells can be found on the ovaries, fallopian tubes, and the tissue lining the pelvis. In rare cases, they may spread beyond the pelvic region. Like the endometrial cells in the uterus, these cells outside the uterus respond to the menstrual cycle. This can cause severe menstrual pain, chronic lower abdominal and pelvic pain, painful intercourse, and fertility issues. The exact cause of endometriosis is not well understood and its diagnosis is often delayed, taking on average 8-10 years from the onset of symptoms. This delay is partly due to the necessity of laparoscopic surgery for definitive diagnosis. Progress in understanding the molecular basis of endometriosis has also been hampered by the invasiveness of the sample collection procedure, as well as the lack of appropriate animal models. Therefore, there is a real need for non-invasive, early diagnosis methods, as well as a better understanding of the disease.

Despite its potential for research and diagnostics, quantitative parameters of MF cellular composition, including cell-count, cell-viability, cell-type representation and transcriptional similarity to endometrium remain only partially described (13,15–17). Moreover, it is unclear whether endometriosis biomarkers identified in other tissue compartments are likewise detectable in MF. To date, the identification of biomarkers for endometriosis has proven challenging due to contradictory results that depend on the exact tissues being contrasted (e.g. peritoneal fluid versus endometriotic lesion versus endometrial biopsy), on the phase of the menstrual cycle at which the tissues were collected, or on the cell-type analysed (see (18) for a systematic review of these issues).

MF can be acceptably and conveniently self-collected by study participants using menstrual cups (19). Here, we profiled self-collected MF in a research setting at single-cell resolution to ask whether MF contains a faithful representation of the cell-type composition and cellular transcriptional states of the endometrium. We then compared the proportion of cell-types and transcriptomes of the different cellular fractions of MF between endometriosis patients and healthy controls.

## Results

### Menstrual fluid collection is a viable sampling alternative to endometrial biopsy

To quantify the cellular and molecular composition of MF, we analyzed 25 self-collected fresh MF samples from 15 healthy women on day 2 of the menstrual cycle (Sup. Table 1). MF samples were serially passed through 100μm and 70μm cell strainers and split into a flow-through and a clump fraction. The retained cellular clumps were gently dissociated prior to further processing. Flow cytometry was performed on each sample fraction staining for CD45+ (immune cells), CD45-EPCAM+ (epithelial) and CD45-EPCAM- (stromal) cell populations (Fig. 1A).

**Figure 1:**
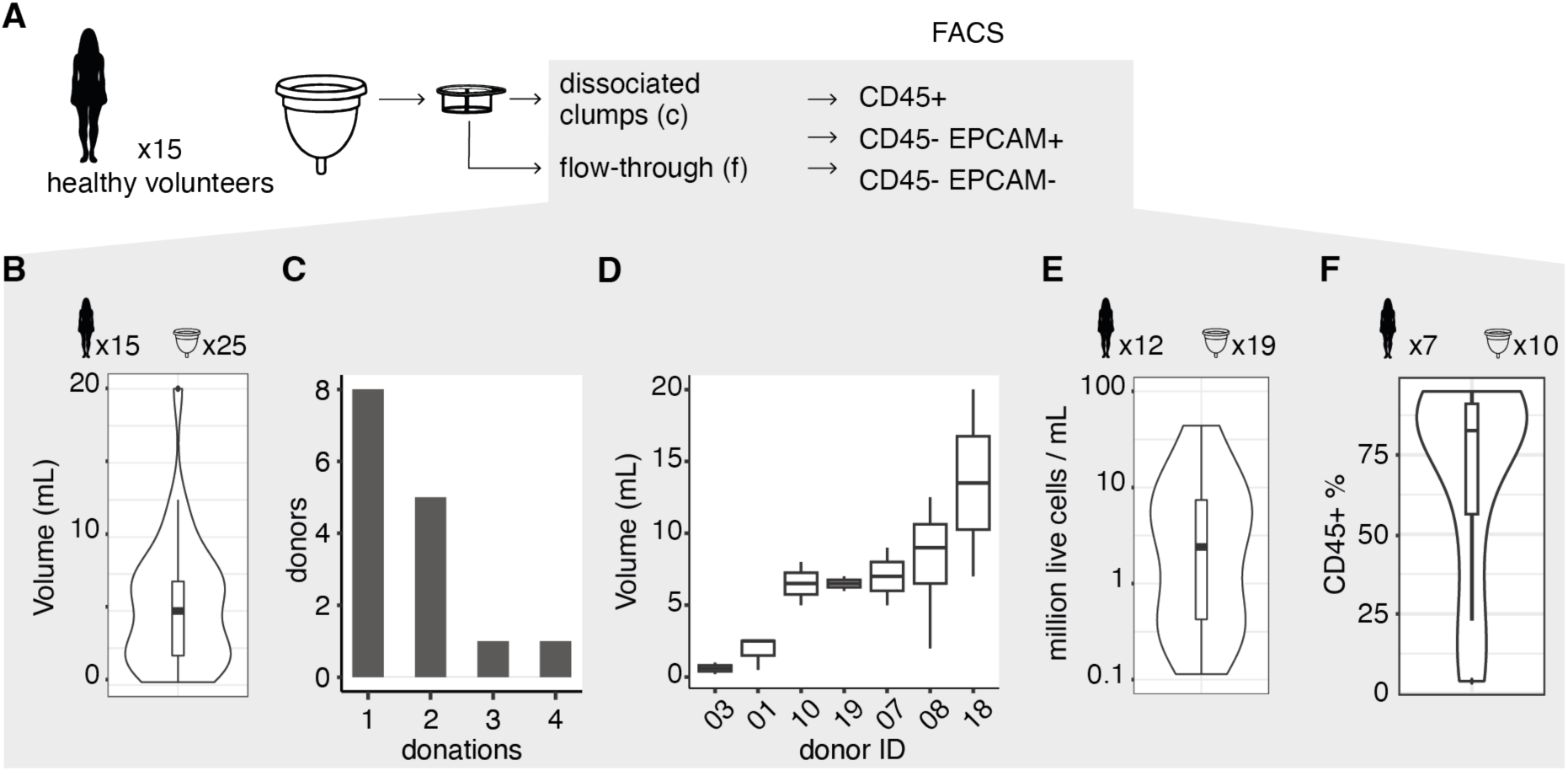
Characterization of menstrual fluid from healthy donors. (A) Sample preparation for the FACS assays. (B) Total sample volumes in mL. (C) Frequency of the number of donations per donor. (D) Volume in mL of MF across 2-4 donations from the same donor. (E) Million of live cells per mL of flow-through. (F) Percentage of CD45+ cells in flow-through.

#### Volume, cell-viability and cell-type composition by fluorescence-activated cell sorting (FACS)

The median volume of MF per sample was 5mL (Fig. 1B), with a potential donor effect (Fig. 1C-D, one-way ANOVA p-value = 0.09), but no age association (generalized linear mixed model with donor random effect p-value = 0.43). Three out of 25 MF samples did not yield any live cells, these samples came from two individuals, both of which subsequently donated again, yielding samples with enough viable cells for processing. The remaining MF samples yielded a median of 2.6 million live cells per mL of flow-through (Fig. 1E). There was no statistical association (Pearson and Spearman correlation coefficient not significantly different from 0 at a 5% significance level) between cell viability and processing time, with the sample with the highest processing time delay after cup removal (12h) containing 63% live cells. After gating out dead cells, flow-through MF was mainly composed of CD45+ cells (Fig. 1F, median 83% of cells).

#### Cell-type composition and comparison to endometrial biopsy by single-cell RNA-seq

To characterize and quantify the cell-types found in MF, we then generated single-cell transcriptomes from 7 MF samples of 4 individuals. For each sample we mixed the 3 sorted populations at a 14/22/64% ratio (Fig. 2A) to obtain a balanced representation of cell-types. Using established marker genes, we identified ciliated, glandular and exhausted epithelial cells, mesenchymal and decidualized stromal cells, smooth muscle/endothelial cells, and immune cells in all 7 samples (Fig. 2B-D, Methods). The ratio of epithelial to stromal and immune cells was similar among MF samples, with epithelial cells comprising around 50% of cells (Fig. 2D).

**Figure 2:**
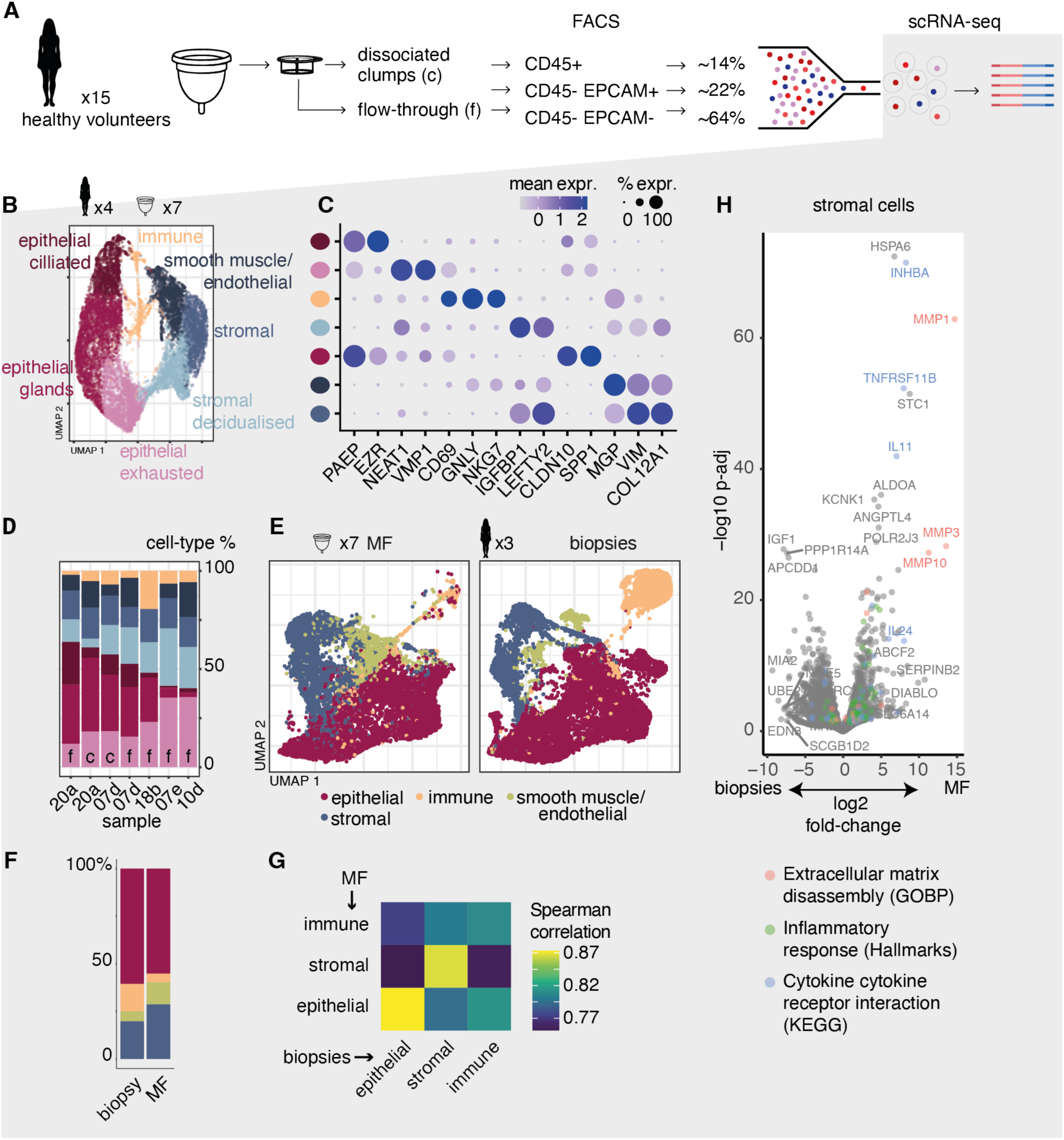
Characterisation of menstrual fluid from healthy donors using single-cell RNA-seq (scRNA-seq). (A) Sample preparation for the scRNA-seq assays. (B) Single-cell UMAP of ciliated, glandular and exhausted epithelial cells, mesenchymal and decidualized stroma, smooth muscle/endothelial and immune cells. (C) Marker genes used for the cell-type annotation. (D) Cell-type abundances in each MF sample (c - clumps, f - flow-through). (E) UMAP of the MF samples integrated with endometrial biopsies from Wang et al. (20). (F) Cell-type abundances in MF and biopsies. (G) Gene expression correlation between the MF and biopsies. (H) Differential gene expression between stromal cells in MF and biopsies. Gene names are shown for the top 15 genes with largest absolute log2 fold changes and for genes with -log10 adjusted p-values greater than 20. Colors indicate if genes are members of the pathways indicated. The pathways chosen were significantly over-represented (FDR q-value <= 0,00027) in gene set enrichment analysis.

We then asked whether the transcriptional profiles of the MF samples correspond to the transcriptional profiles of freshly isolated endometrial cells by computationally integrating our single-cell data with single-cells from 3 endometrial biopsies (20) in the late-secretory and proliferative phases of the menstrual cycle (Fig. 2E-F). Transcriptionally, MF epithelial, stromal and immune cells were more similar to their corresponding cell-types in endometrial biopsies than to one another, indicating that these cells are able to retain their cellular identity despite being shed (Fig. 2G). Epithelial and stromal cells exhibited the highest degree of transcriptomic concordance between MF and endometrial biopsy cells, whereas the immune cells were more divergent, indicating that immune cells are more sensitive to shedding or experimental manipulation.

Despite the overall transcriptomic concordance, we expected expression differences between MF and the endometrial biopsies to reflect differences in cycle stage. The endometrial biopsy samples used came from cycle stages with high progesterone levels, which represses, among others, expression of many matrix metalloproteinases (MMPs). Progesterone withdrawal at menstruation triggers expression of MMPs (21), an effect mediated by cytokines including tumor necrosis factor-α (TNF-α). In addition, a large number of neutrophils loaded with MMPs are recruited through induction of chemokines such as interleukin 8 (CXCL8) (21). Differential gene expression comparison between MF and the endometrial biopsy stromal cells revealed much higher expression in MF of MMPs, members of the TNF-α signaling pathway and *CXCL8* (Fig. 2H, Sup. Table 2).

In sum, our data show that sampling self-collected MF is a viable strategy for obtaining large numbers of live epithelial, stromal and immune cells from the human endometrium in an outpatient setting without prior scheduling. MF samples capture the main cell types of the endometrium and are transcriptionally similar to cells collected via invasive biopsy.

### Properties of MF from patients with endometriosis and healthy controls

To assess the potential of MF to provide insight into female reproductive tract disorders, we collected 36 fresh MF samples from 7 women with a confirmed diagnosis of endometriosis and 11 age-matched healthy volunteers (Fig. 3A, Table 1, Sup. Table 3). Samples were processed by density gradient to remove red blood cells. High volume samples were split into CD45+ and CD45-fractions by MACS. For a subset of samples, we used FACS to estimate proportions of CD45+ vs. CD45-cells and the CD45-fraction was further sorted into CD105+/- cells for a estimation of the fraction of putative mesenchymal stem cells present (22).

**Figure 3:**
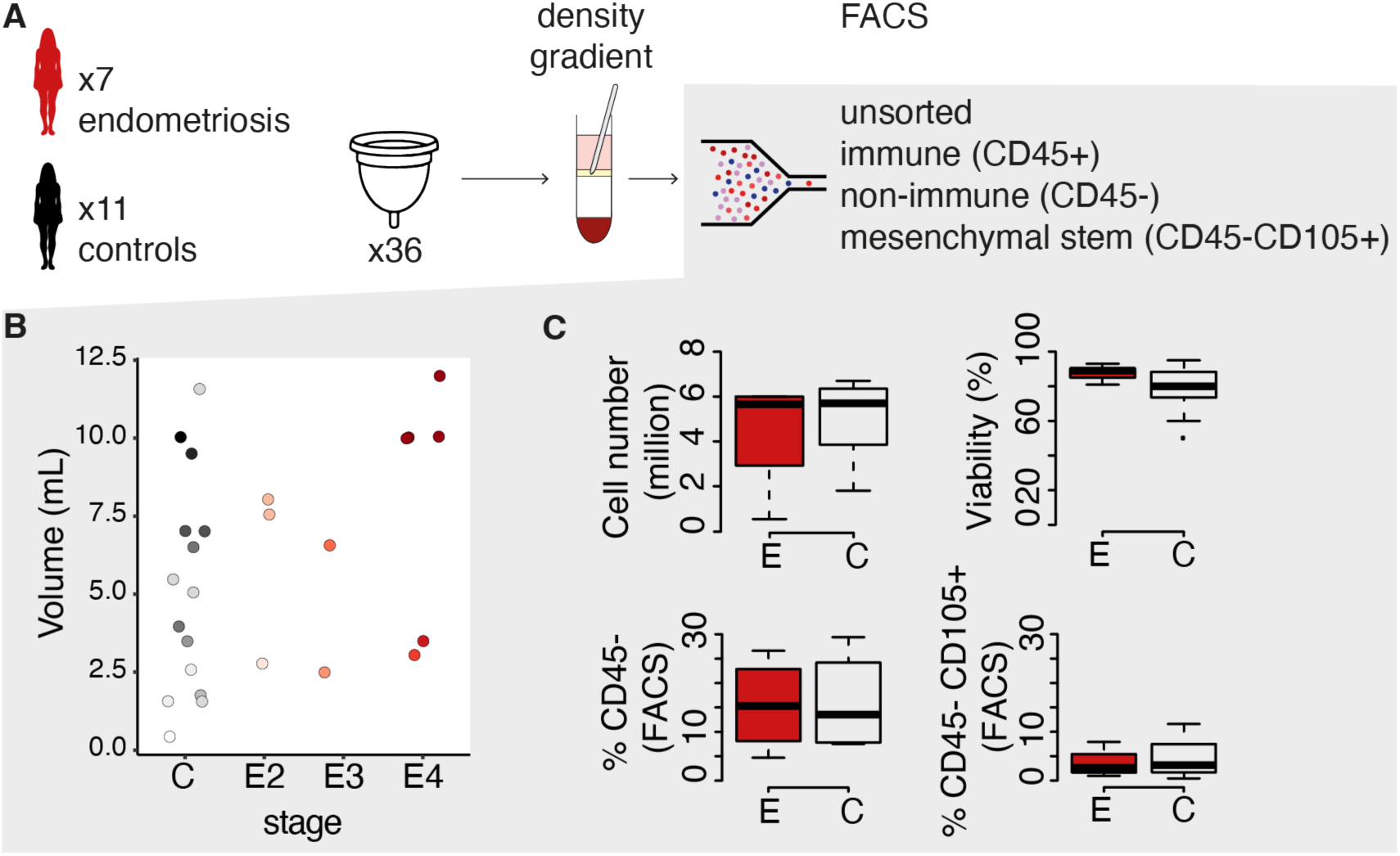
Comparison of menstrual fluid volume, cell number and cell-type proportions between endometriosis patients and healthy controls. (A) Sample preparation procedure. (B) MF volume in controls - C, and endometriosis stages 2 to 4 (E2-E4). Points are coloured by donor. (C) Cell number, viability, fraction of CD45+ cells and fraction of CD45+CD105+ cells in endometriosis - E, and controls - C.

**Table 1:**
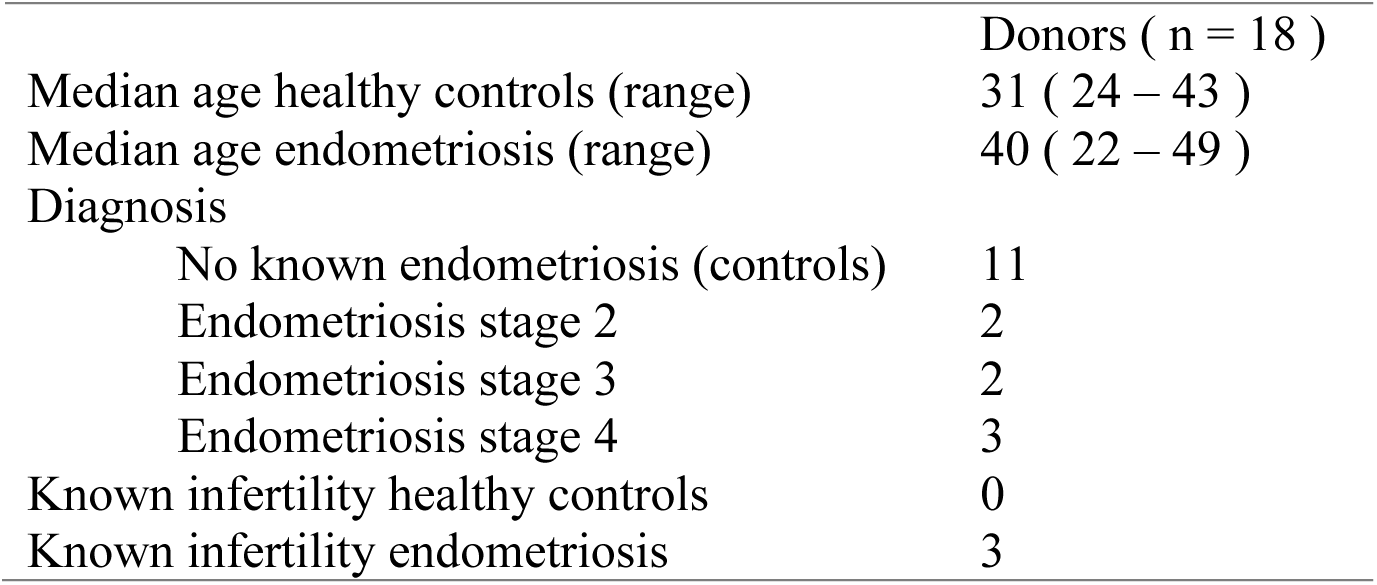
Demographic and disease characteristics of the endometriosis and healthy controls.

Once more, we observed a donor effect on volume (Fig. 3B, ANOVA p-value 0.054). However, we did not observe any statistically significant differences between endometriosis and healthy volunteer-derived MF samples in volume, cell number, cell viability, fraction of CD45+/- cells, or fraction of mesenchymal stem cells (CD45-CD105+) (Fig. 3B,C). Moreover, MF volume was not statistically significantly different between endometriosis stages at this sample size (Fig. 3B). The vast majority of samples could be successfully cultured over multiple passages, under standard mesenchymal cell culture conditions (Methods), irrespective of whether they came from cases or controls.

#### Cell-type composition of ex vivo and in vitro MF cells by RNA-seq

To molecularly characterize both freshly isolated (*ex vivo*) and cultured (*in vitro*) samples, as well as identify putative differences between endometriosis patients, we performed 3’ bulk RNA-sequencing on 93 samples (Fig. 4A). A principal component analysis of gene expression patterns revealed two main sample groups: samples that were processed directly after isolation (*ex vivo*) and those that were cultured (*in vitro*) (Fig. 4B). An over-representation analysis of genes negatively correlated with PC1 (i.e. genes with higher expression in *ex vivo* samples) revealed these to be highly enriched for blood cell-type markers, i.e. monocytes, dendritic and macrophage-specific genes (Fig. 4C, Sup. Table 4). Cultured samples were transcriptionally more homogeneous than freshly isolated samples, likely due culture-induced selection for CD45-cells. Indeed, we observed that the CD45+ cell fraction typically failed to attach, in line with previous reports that have shown preferential attachment of menstrual stromal cells (so-called menstrual-derived stem cells, or colony-forming cells) upon culture (23). Nevertheless, when comparing *in vitro* cells to CD45-*ex vivo* cells, we observed that the transcriptional profiles remain distinct, suggesting major transcriptional shifts induced by cell culture (Fig. 4D, Sup. Table 5). In contrast, *ex vivo* CD45+ samples showed higher similarity to *ex vivo* non-sorted (*unsorted*) samples (Fig. 4E), in line with the observation that most MF cells are CD45+ as determined by FACS.

**Figure 4:**
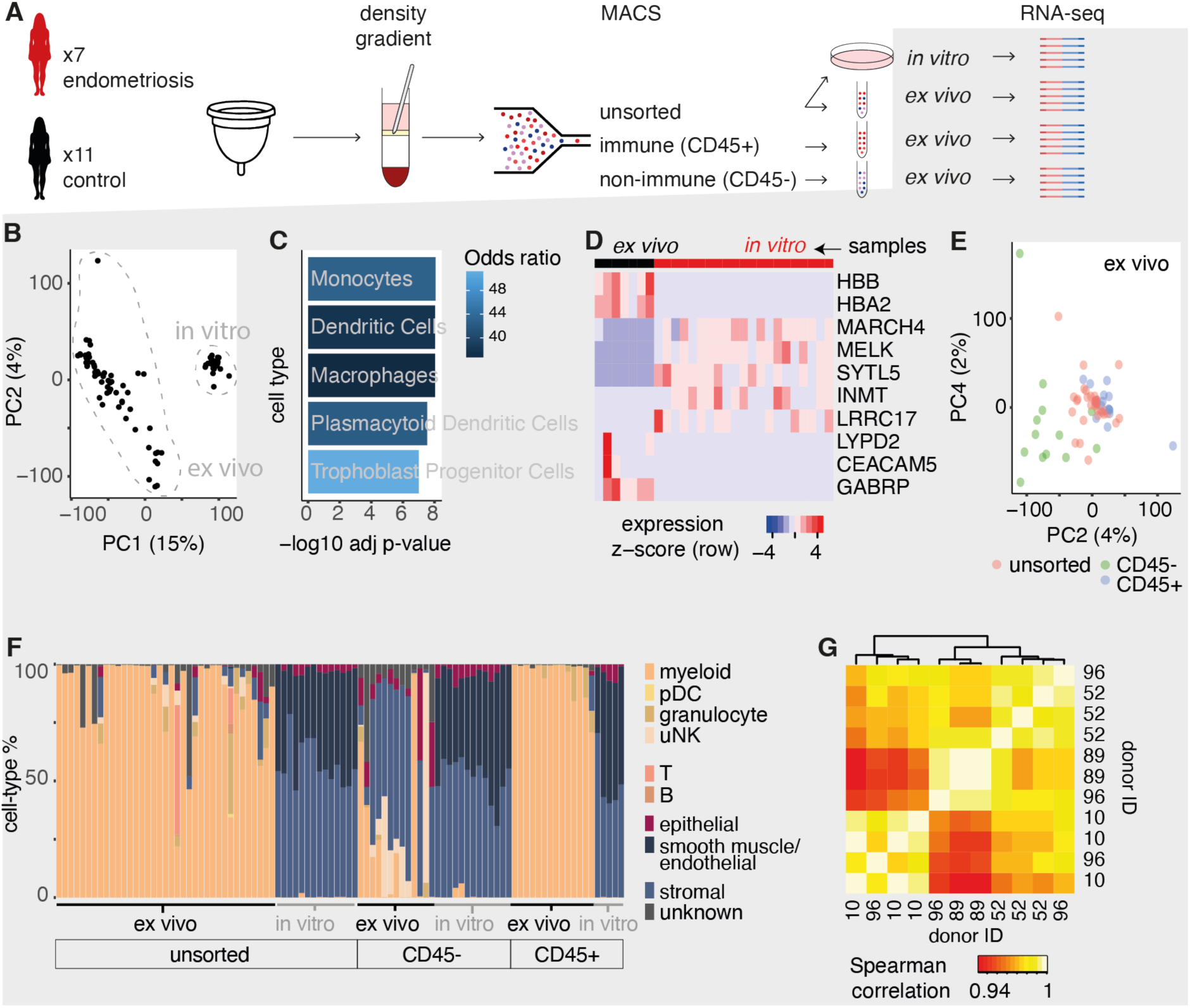
Cell-type composition of MF samples by cell-type enrichment method. (A) Sample preparation procedure. (B) Principal component analysis of all samples using the full transcriptome. Shown in parentheses on the axes is the percentage of variance explained by each of the principal components. (C) Over-representation analysis of the top 30 genes positively associated with PC1 in the cell-type marker gene-sets from PanglaoDB(54). P-values were calculated using Fisher’s exact test and corrected for multiple testing using the Benjamini-Hochberg procedure. (D) Gene expression heatmap of the 10 most significantly differentially expressed genes between CD45-*ex vivo* and all *in vitro* samples. (E) Principal component analysis of *ex vivo* samples. (F) *In silico* cell-type deconvolution of the bulk RNA-seq samples using MuSiC. pDC - plasmacytoid dendritic cells, uNK – uterine NK cells. (G) Spearman correlation of the whole CD45+ *ex vivo* transcriptomes for donors that donated more than once. The row-side dendrogram (identical to the column-side one) was omitted.

To confirm the differences in cell-type composition, we used an *in silico* transcriptome deconvolution method (24) and a single-cell reference dataset (13) to estimate cell-type fractions from our bulk RNA-seq samples. We found that indeed *in vitro* samples are preponderantly composed of stromal, smooth muscle and/or endothelial-like cells, whereas *ex vivo* samples are preponderantly composed of myeloid cells (Fig. 4F).

In sum, MF collected *ex vivo* and *in vitro* display transcriptional changes consistent with changes in the cell-type enriched, as well as a culture induced phenotype. *In vitro* samples only capture a subset of the cell populations initially present in MF, preponderantly stromal-like cells.

#### Donor effect on transcriptomic profiles

Cycle-to-cycle variability in MF transcriptomic profiles has not been previously characterised. There is a concern that samples might be too variable due to unpredictability in tissue breakdown and exact timing of sample collection. Leveraging on having multiple longitudinal samples from the same donor (Fig. 3B), we asked if MF samples are transcriptionally stable across cycles of the same donor. To do so, we looked at transcriptomic concordance among CD45+ *ex vivo* samples, as these were the most homogeneous in terms of their composition (Fig. 4F). Samples from the same donor clustered together more often than expected by chance, suggesting that donor effects can be observed over multiple cycles (Fig. 4G).

### Identification of potential transcriptional biomarkers of endometriosis in MF

To identify MF gene expression markers that might distinguish endometriosis from healthy controls, we used three different statistical models on various subsets of the 93 RNA-seq assays performed. First, we identified endometriosis biomarkers that are detectable in stromal-like CD45-cells. Second, we identified biomarkers that are detectable in immune CD45+ cells. Finally, we implemented a model to identify biomarkers that are detectable irrespective of cell-type enriched or whether the sample is freshly isolated or cultured.

#### Differentially expressed genes in CD45-stromal-like cells

In this model, we sought to identify differentially expressed genes that can be found in *in vitro* CD45-samples. We found 256 genes differentially expressed at a 5% false discovery rate (Fig. 5A, Sup. Table 6). These included the down-regulation of stromal decidualization genes (20,25–27) in endometriosis patients when compared to healthy controls, supporting findings that decidualization is compromised in cultured (28,29) and freshly isolated stromal cells from endometriosis patients (11,13,14). Likewise, among genes downregulated in endometriosis patients we found an enrichment for targets of estrogen receptor alpha (*ESR1*), whose expression has been previously found to be suppressed in endometric stromal cells (30). Additionally, we found that inflammation related pathways (TNFa signaling and IL-2/STAT5), were down-regulated in endometriosis patients, whereas epithelial to mesenchymal transition (EMT) and angiogenesis were upregulated (Sup. Table 7).

**Figure 5:**
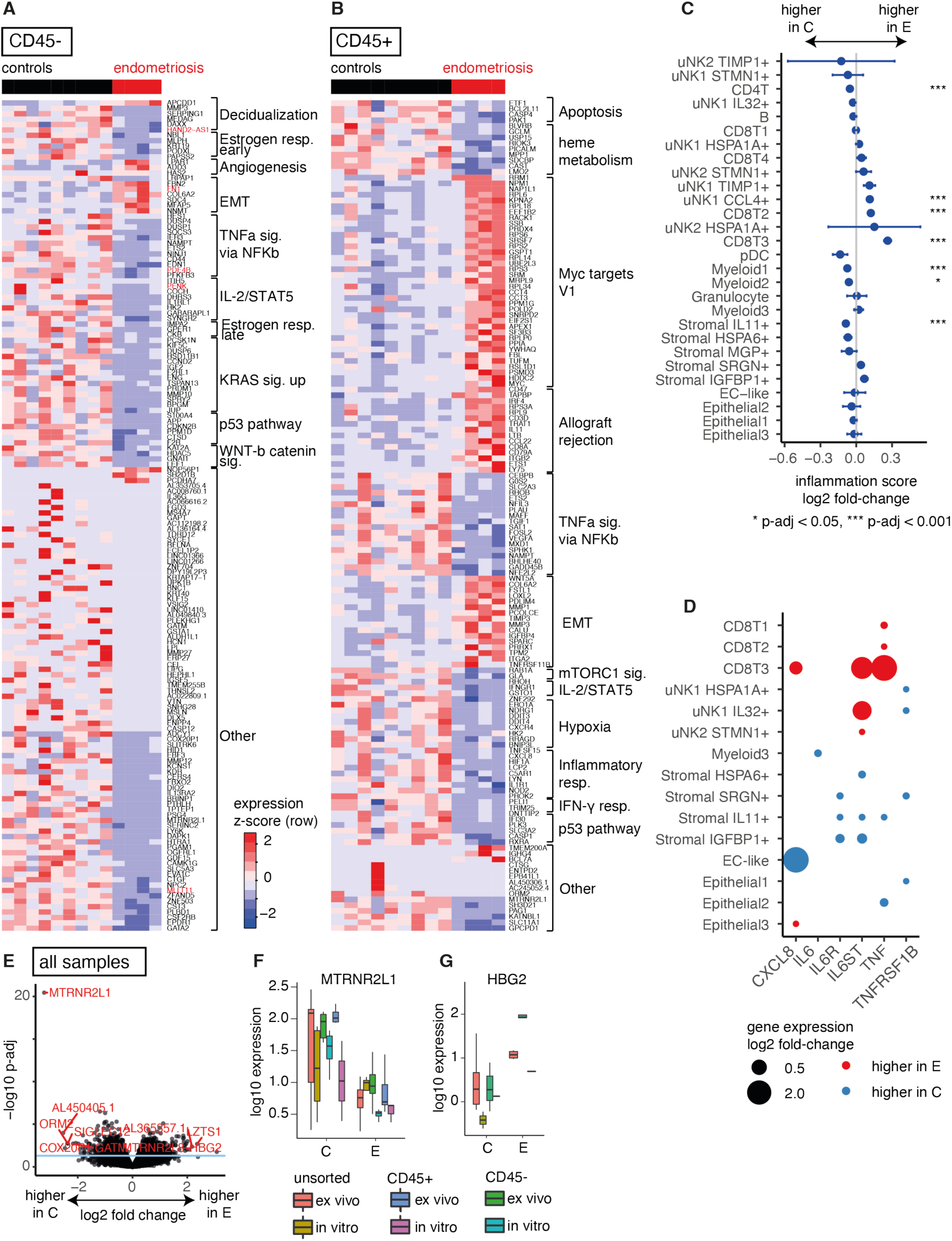
Comparison of the transcriptomes between endometriosis patients and healthy controls. (A) Gene expression heatmap of 150 genes differentially expressed genes in CD45-*in vitro* samples (columns). The genes shown are either 1) members of pathways significantly enriched among differentially expressed genes, or 2) top most differentially expressed genes according to p-value and absolute log2 fold-change (“Other”). (B) Gene expression heatmap of 150 genes differentially expressed genes in CD45+ *ex vivo* samples (columns). The genes shown are either 1) members of pathways significantly enriched among differentially expressed genes, or 2) top most differentially expressed genes according to p-value and absolute log2 fold change (“Other”). (C) Log2 fold-change of the inflammatory signaling pathway activity between endometriosis patients – E, and controls – C in the different cell-types defined in Shih. et al. Horizontal bars show standard errors and asterisks indicate significance level of a t-test performed for each cell-type independently. P-values were adjusted for multiple testing with the method from Benjamini-Hochberg. (D) Log2 fold-change of the expression of selected ligand-receptors involved in inflammatory pathways between endometriosis – E and controls – C. (E) Volcano plot of the differential expression of endometriosis - E - versus healthy controls – C using all available samples. Points above the horizontal blue line indicate genes with multiple-testing adjusted p-values below 0.05. (F) Normalised expression levels of MTRNR2L1 in endometriosis - E and control - C samples. (G) Normalised expression levels of HBG2 in endometriosis - E and control - C samples.

Among down-regulated genes we found multiple endometriosis previously implicated in the disease including *PENK*, *MLLT11*, *HAND2-AS1* and *PDE4B*.

The endogenous opioid peptide precursor proenkephalin (*PENK*) has been previously reported to be up-regulated in whole-endometrium (31,32) of endometriosis patients when compared to healthy controls. In contrast, *PENK* expression was found to be down-regulated in ectopic stroma when compared to eutopic stroma of patients (33). The discrepancy in the direction of regulation may have to do with the specific cell-types assayed, as well as the cycle phase at which endometrial samples were obtained, as *PENK* expression is modulated by the menstrual cycle (31). Interestingly, a decrease in opioid peptides has been previously suggested to be involved in the maintenance of chronic inflammation in endometriosis (34).

Consistent with our findings, the expression of mixed-lineage leukemia translocated to 11 (*MLLT11*) was previously found to be reduced in the ectopic stromal cells of women with advanced endometriosis compared to healthy controls, and its down-regulation found to be associated with an increased stromal cell adhesion phenotype (35).

HAND2 Antisense RNA 1 (*HAND2-AS1*), whose expression is coordinated with *HAND2* in human endometrial stromal cells, was also previously found to be reduced in ectopic lesions from patients compared controls, primarily expressed by stromal cells as opposed to epithelial, and that its silencing impairs stromal cell decidualization (20,26,27).

Finally, genetic polymorphisms in cAMP-specific 3’,5’-cyclic phosphodiesterase 4B (*PDE4B*) were recently proposed to be implicated in the etiology of endometriosis (36).

Relatively fewer genes were found to be up-regulated in endometriosis patients when compared to controls. Notably, the most significant up-regulated gene was fibronectin 1 (*FN1*). A genome-wide meta-analysis and a subsequent targeted study have implicated single nucleotide polymorphisms in FN1 with moderate to severe endometriosis (37,38), and relaxed fibronectin is being investigated as a potential target for imaging endometriotic lesions (39). At the protein level, studies comparing eutopic and ectopic endometrium from patients have yielded contradictory results (39). However, fibronectin levels in plasma and peritoneal fluid have consistently reported elevated levels in endometriosis patients when compared to healthy women (40,41).

#### Differentially expressed genes in CD45+ immune cells

We found 792 differentially expressed genes among the CD45+ *ex vivo* samples (Fig. 5B, Sup. Table 8, the one sample with an abnormally high percentage of stromal cells in Fig. 4D was excluded). We found a trend for lower expression of immune-related genes in CD45+ MF samples originating from endometriosis patients, in particular for genes related to inflammatory pathways (TNFa signaling, Inflammatory response, IFN-gamma response, Sup. Table 9).

This was intriguing, as the production of potentially pro-inflammatory cytokines, such as IL-6, has been previously found to be up-regulated in cultured and fresh stromal cells from ectopic and eutopic endometrium from patients compared to controls (13,42). To explore the activation of inflammatory pathways in specific cell-types from endometriosis patients and controls, we used AUCell (43) to score the activity of the MsigDB “inflammatory response” pathway in each individual cell in the dataset from Shih et al. (13) (Methods).

In agreement with our findings, we confirmed that the activity of the inflammatory signaling pathway was significantly downregulated in myeloid (the most abundance cell-type in CD45+ *ex vivo* samples) and CD4+ T cells, and in a subtype of stromal cells of endometriosis patients. In contrast, the pathway was upregulated in uNK cells and two subtypes of CD8+ T cells of endometriosis patients (Fig. 5C). A similar pattern emerged when looking at the expression of specific inflammatory ligand receptors (Fig. 5D).

At the opposite end of the spectrum, we identified several genes significantly more highly expressed in endometriosis patients versus healthy controls, many of which had also previously shown to have increased expression in the eutopic endometrium of endometriosis patients compared to healthy controls (e.g. *CALD1* (44) and *SEMA3A* (45), which has been proposed to be involved in macrophage recruitment to endometriotic lesions (46)), or increased expression in the eutopic endometrium of severe compared to mild endometriosis patients (*TEAD2* (47)). Unlike the downregulated genes, these genes were typically not immune-cell specific, but rather connected to tissue invasiveness, epithelial to mesenchymal transition or myc targets (Fig. 5B). C-myc expression has been previously reported to be altered in the eutopic endometrium of endometriosis patients when compared to controls which, together with a reduction in cell death by apoptosis, is thought to play a role in facilitating the invasive features of endometriotic endometrium (48).

Interestingly, among immune-related genes, endometriosis patients also had significantly higher expression of *CD8A* (Fig. 5B). The abundance and activation levels of CD8 T cells has been a topic of considerable interest in endometriosis research. However, results vary depending on the tissue analysed (49). Our study supports the notion that CD8+ T cells (or subsets thereof (13)) are enriched in the menstrual fluid of endometriosis patients.

#### Differentially expressed genes in all cell and sample types

In the model designed to identify biomarkers that are detectable irrespective of cell type enriched or whether the sample is freshly isolated or cultured, we identified a large number of differentially expressed genes (Fig. 5F, Sup. Table 10). Many of these genes, such as *MTRNR2L1* and *HBG2*, were not, to our knowledge, previously described as biomarkers of endometriosis.

*MTRNR2L1* was detected by all 3 models as being substantially upregulated in healthy controls compared to endometriosis patients (Fig. 5G). As high *MTRNR2L1* expression in human endometrium is highly specific to endometrial epithelial glandular cells (50), the lower expression of this gene in endometriosis patients might be driven either by lower expression levels in their epithelial cells, or by a proportion of epithelial cells present in their MF. Interestingly, despite not having been previously linked to endometriosis, *MTRNR2L1* was found to be downregulated in the eutopic endometrium of adenomyosis patients compared to healthy controls (51).

*HBG2* is more highly expressed in endometriosis patients than in controls (Fig. 5H). As high expression of *HBG2* is very highly specific to macrophages and erythroid cells and lowly expressed in all cell-types of the endometrium (50), we hypothesize that the origin of this signal may be in a higher proportion of macrophage cells in endometriosis patients, or in a higher amount of ambient RNA from higher levels of cell death in the samples of endometriosis patients. Intriguingly, *HBG2* was previously found to be over-expressed in the menstrual endometrium of women with heavy menstrual bleeding when compared to controls (52).

## Discussion

This study demonstrates the feasibility of using self-collected MF to obtain a comprehensive cellular and molecular snapshot of the human endometrium. Our data show that MF collection is not only viable but also yields a substantial number of live cells, including epithelial, stromal, and immune cells. The similarity in transcriptional profiles between MF and endometrial biopsies and recapitulation of menstrual cycle dynamics reinforces the reliability of MF as a representative sample of the endometrial environment. This representativeness opens avenues for exploring menstrual fluid in research settings and potential clinical diagnostics.

Our pilot study reports on useful parameters for future large-scale studies. Our data show that MF is primarily composed by CD45+ myeloid cells, which dominate the transcriptomic signal. Unsorted *ex vivo* samples vary widely in cell-type composition with no association with disease state, highlighting the importance of enriching for the target cell-type in both bulk or single-cell transcriptomics assays. We report differences between *ex vivo* and *in vitro* (cultured cells) cells, with the former containing a large number of immune-related cells, and the latter being composed mainly of stromal-like cells. This stresses the importance of probing fresh cells directly or having alternative specialized culture conditions if the immune compartment is to be assessed. We also report on genes whose transcription in stromal cells has been previously reported to be dysregulated in endometriosis patients, but which we found to be affected by cell culture (e.g. *MARCH4*, *INMT* or *GABRP* (32,53)) or menstrual cycle stage (18) (e.g. *PENK*).

In patients with endometriosis, our MF analysis revealed distinctive gene expression patterns. Notably, the differential expression of genes such as *FN1*, *PDE4B*, *PENK*, *MLLT11* and *HAND2-AS1* in stromal-like cells of endometriosis patients aligns with previous findings and underscores their potential roles in the disease’s pathogenesis. Furthermore, the dysregulation of immune-related genes in endometriosis patients suggests an altered immune landscape in endometriosis, possibly contributing to the disease’s inflammatory component. The identification of *MTRNR2L1* and *HBG2* as potential biomarkers that are robust to cell culture and cell-type enrichment is intriguing. These findings necessitate further research to validate and understand the roles of these biomarkers in endometriosis.

The use of MF for diagnosing endometriosis offers a promising non-invasive alternative to current diagnostic methods, which are often invasive and delayed. Our pilot study was limited by a relatively small sample size. Further research with larger cohorts with standardized collection procedures is essential to validate our findings and establish MF-based diagnostics in clinical practice.

In conclusion, our study highlights the potential of menstrual fluid as a rich, non-invasive source for studying the endometrium and diagnosing endometriosis. This work paves the way for further studies to explore the full diagnostic potential of menstrual fluid and its application in women’s health. Combined with banking stem cells isolated from MF, MF sampling can enable powerful functional studies of gynecological disease.

## Methods

### Sample collection for the healthy donor cohort

The study was approved by the South West - Frenchay Research Ethics Committee (IRAS project ID 221351). Fifteen healthy participants (menstruating, self-defined healthy females with no other selection criteria) were recruited from employees, visitors and students at the Wellcome Trust Sanger Institute. All samples and data collected were solely identified by a randomly assigned identifier and the research team had no access to the participant names. No links were kept between A) the consent forms containing the participant name and B) the samples and sample associated data collected. Participants were between 24 and 47 years old with self-reported regular menstrual cycles. Two of the 15 participants were users of combined oral contraceptive pills, and a third one of a contraceptive implant. All women were requested to collect their complete menstrual fluid during day 2 of their period using a silicone menstrual cup (Mooncup®). These cups were purchased by the investigation team for the purpose of this investigation. We do not hold any commercial link with this provider. Size A was recommended if the person was aged 30 and over and/or has given birth vaginally at any age. Size B was recommended if the person was under 30 and had not given birth vaginally. Participants were asked to pour and/or pipette the sample from the Mooncup into a sample collection tube and to deliver the sample tube to the study collection boxes within 6 hours of menstrual cup removal. Donors were able to donate samples up to 6 times each and at each donation filled in a questionnaire with demographic and biologic data (age, time of cup removal, current use of contraception, medication and ongoing illness). All sample collection tubes and questionnaires within a kit were labeled with the same randomly assigned identifier, thereby completely anonymising the samples but allowing multiple donations from the same participant to be linked. Sample delivery was unscheduled and samples were processed upon receipt from the collection boxes (within 30m to 12h from the time of menstrual cup removal, of this time up to 3h at RT and the remainder in the fridge at 5°C).

### Sample processing for the healthy donor cohort

The total volume, appearance (color, haemolysis and viscosity scores) and presence of clumps/mucus were assessed for each sample. Five of the samples had high mucous content and were centrifuged at 300g for 5 minutes. The supernatant was transferred to a new tube, spun at 2000g for 3 minutes and resuspended with 50mL PBS. All samples were then serially filtered with 100μm and 70μm nylon mesh sieves (Falcon). Clumps were backwashed from the sieves with PBS and digested with collagenase for 20–60 minutes at 37°C. Flowthrough and clumps were briefly centrifuged and subject to red blood cell lysis (Biolegend Inc.). Cells were counted with Trypan blue. Samples with more than 1 million live cells were processed by flow cytometry.

### Flow cytometry staining and acquisition for the healthy donor cohort

After digestion, cells were washed and centrifuged at 2000rpm for 3 minutes. Cells were resuspended in PBS plus Human TruStain FcX Blocking solution (BioLegend) and incubated for 5 minutes at room temperature in the dark. 5μl/million cells of CD45 (clone 2D1, BioLegend) and EPCAM (clone 3C4, BioLegend) antibodies were added and incubated for 1h at 4°C in the dark. Post staining, cells were washed and centrifuged at 2000rpm for 3 minutes. 1μl/mL of DAPI was added and incubated for 5 minutes at RT. Cells were washed twice, filtered with a 50μm FACS filter and analyzed using SH800S, MoFlo XDP or BD Influx cell sorters, depending on instrument availability, according to manufacturer’s instructions and gating strategies. After sorting, the CD45+, CD45-EPCAM- and EPCAM+ fractions were collected for 10x single-cell RNA-sequencing.

### Generation of single cell transcriptomes

Single-cell suspensions were loaded per channel of ChromiumTM Single Cell Chips (10X Genomics® Chromium Single Cell 3’ Reagent Kits v2.0), aiming for a recovery of 4,600 cells. Reverse transcription and library construction were carried out according to the manufacturer’s recommendations. Libraries were sequenced on an Illumina HiSeq 4000 using paired-end runs of 150bp.

### Computational analysis of the single-cell RNA-seq data

Raw sequencing reads were processed using the Cell Ranger analysis pipeline (55). The “cellranger count” command was used to generate filtered and raw matrices. Reads were aligned against the human genome version GRCh38. Raw gene barcode count matrices were processed using CellBender for unsupervised denoising (56) and further analyzed using the R package Seurat (57). To remove low quality cells, an adaptive filtering threshold approach was used based on extreme numbers of counts (count depth) and extreme numbers of genes per barcode. Cells were filtered when counts or genes per barcode were less than 99% of all cells, or when the mitochondrial content was higher than 10%.

To annotate cell types, we used two strategies: annotation of individually processed samples, and annotation of all samples integrated together. For the individually processed samples, counts were normalized using the SCT normalization approach of Seurat. Sample-specific UMAPs were constructed using a subset of genes exhibiting high cell-to-cell variation which were identified by modeling the mean-variance relationship. The top 3000 features were used to perform PCA analysis. To cluster the cells, a K-nearest neighbor (kNN) graph based on the euclidean distance in PCA space was first constructed using the first 30 PC components as input. Next, the Louvain algorithm was applied to iteratively group cells. We identified the cell types in each cluster using a combination of manual and automated approaches using known marker genes (Fig. 1E). First, clusters were assigned to known cell populations using cell type– specific markers obtained through the FindAllMarkers function. Multiple testing correction was performed using Benjamini-Hochberg procedure. Second, the R package Garnett (58) in cluster extension mode was used to annotate cells in a semi-automated manner. Integration of samples was performed using reference-based canonical correlation analysis together with SCT normalization (Seurat), and marker based annotation was performed as above. A consensus of the three annotation strategies was used to annotate each sample.

Integration of the MF samples with the endometrial biopsies was performed using reference-based canonical correlation analysis together with SCT normalization (Seurat) using 3000 integration features.

Differential expression analysis of menstrual fluid versus the endometrial biopsies was performed by generating pseudo-bulks for each of the main cell-types: stromal, epithelial and immune. Pseudo-bulking was performed by summing up the counts of all cells of each cell-type by sample. Only genes that had non-zero counts in at least one MF and one biopsy sample were kept. Samples were then used as replicates in a DESeq2 analysis using default parameters. Log fold change shrinkage using the lfcShrink function with apeglm was performed on the results.

Gene set enrichment analyses were performed using the GSEA tool (v4.3.3, (59)). Sorted differentially expressed genes by log fold change were used as input to a weighted analysis using the following gene-sets: Hallmarks, GO Biological Process, Reactome and KEGG.

### Sample collection and processing for the endometriosis cohort

We recruited age-matched (20–45) healthy female volunteers and endometriosis patients through advertising at endometriosis support groups as well as the EPFL campus. The study received ethical approval by the Swiss Cantonal Authorities Vaud, CER-VD, project nr. 2016-00770. All samples and data collected were solely identified by a randomly assigned identifier. Samples were processed within 6h by a standard density gradient-based cell isolation protocol (Histopaque). The viability and the number of nucleated cells in the cell suspension was determined using a Nucleostainer and when possible, further processed by MACS into a CD45+ and CD45-fraction. For a subset of samples, surface marker gene expression was determined by FACS. The isolated single cell suspension was diluted to 1 × 10^7^ cells/ml with FACS buffer (DPBS −/−) with 1% human platelet serum and the following fluorophore-conjugated antibodies were added: anti-human CD45 for identifying hematopoietic cells, and anti-human CD105 for identifying mesenchymal stromal cells. 7-Aminoactinomycin D (7-AAD) was used for assessing viability and Syto40 was used for discerning nucleated cells. Cell culture was performed using high glucose MEMalpha medium supplemented with 5% platelet serum and 50 ug/ml Primocin. TrypLE Select reagent was used to collect the cells from the cell culture plates.

### Generation of bulk RNA-seq for the endometriosis cohort

RNA was collected from fresh or cultured samples into Tri-Reagent and bulk RNA-seq was performed as previously described, using BRB-seq, a highly multiplexed 3’ end protocol (60).

### Computational analysis of the RNA-seq for the endometriosis cohort

Sequencing results were processed in the Deplancke laboratory by a standard in-house pipeline, consistent with published methodology to obtain digital gene expression values for each cell and estimated expression for each sample. In brief, sequenced tags were demultiplexed (at sample and library level) and fastq files containing 62-bp-long single-end sequenced tags (reads) were were trimmed and filtered using prinseq 0.20.3 and cutadapt 1.5 and subsequently aligned to the Ensembl 84 gene annotation of the hg19 human genome using STAR 2.4.0g. The number of tags per gene was calculated using htseq-count 0.6.0 with the parameters ‘htseq-count -m intersection-nonempty -s no -a 10 -t exon -i gene_id’.

The over-representation analysis in Fig. 4C was performed by taking the top 30 genes most anticorrelated with PC1 from Fig. 4B and testing their enrichment in the cell-type marker gene sets from PanglaoDB (54), using the hypergeometric test implemented in Enrichr (61).

The decomposition of the bulk-RNA-seq data into cell-type compositions was performed using raw counts and cell-type annotations from the dataset in Shih et al.(13) as reference, and the sample deconvolution method implemented in the MuSiC R package (24), with default parameters.

The differential gene expression analyses of endometriosis versus healthy controls were performed using the standard DESeq2 workflow (62). For the CD45-*in vitro* and CD45+ *ex vivo* cell analyses we used endometriosis (yes or no) as a fixed effect. For the “all cell and sample types” analyses we used type (*ex vivo* or *in vitro*), cell-type (unsorted, CD45+ or CD45-) and endometriosis (yes or no) as fixed effect terms.

Over representation analysis for Fig. 5A-B was performed by taking all genes significantly up or down regulated in each analysis and testing for their enrichment in the MSigDB Hallmarks gene sets (63) using the hypergeometric test implemented in Enrichr (61) and correcting for multiple testing. Additionally, in Fig. 5A we included differentially expressed decidualization-associated genes. We defined decidualization-associated genes as genes whose expression peaks in stromal cells during decidualization in Wang et al. (20) or that are known from the literature.

For the scoring of the “Inflammatory signaling” pathway in the single-cells from Shih et al.(13), we downloaded the gene names for this pathway from MSigDB (63) and used AUCell (43) with maxrank set to 800. To test for significance of differential scoring between cell-types we transformed the scores with log2(score+0.0000001) and then used a linear model with endometriosis status (yes or no) as predictor.

## Data Availability

All data produced in the present work are contained in the manuscript and available online at 10.5281/zenodo.11105267

https://zenodo.org/records/11105267

## Supplementary Materials

Sup. Table 1: Donor and sample characteristics for the healthy single-cell cohort.

Sup. Table 2: Differentially expressed genes between MF and endometrial biopsies.

Sup. Table 3: Donor and sample characteristics for the endometriosis and controls bulk-RNA-seq cohort.

Sup. Table 4: Over-representation analysis of in vitro associated genes in the PanglaoDB cell-type marker gene sets.

Sup. Table 5: Differentially expressed genes between in vitro and ex vivo CD45-samples.

Sup. Table 6: Differentially expressed genes between CD45-in vitro samples of endometriosis patients versus controls.

Sup. Table 7: Over-representation analysis of the significant (padj < 0.05) genes from Sup. Table 6.

Sup. Table 8: Differentially expressed genes between CD45+ ex vivo samples of endometriosis patients versus controls.

Sup. Table 9: Over-representation analysis of the significant (padj < 0.05) genes from Sup. Table 8.

Sup. Table 10: Differentially expressed genes between all samples of endometriosis patients versus controls.

## Author contributions

AG and PCS: designed the study.

GC: provided critical input on study design and volunteer recruitment.

CB, JR, MB, MZ and DA: collected, processed samples and performed experiments. AG, PCS and EA: analysed data.

AG: wrote the manuscript.

BD: provided resources and input on data interpretation. PCS: edited the manuscript.

All authors: read, commented and approved the manuscript.

## Acknowledgements

This work was supported by an Early Detection Gynaecological Cancers Pump Priming award by CRUK to AG. PCS was supported by an HFSP Postdoctoral Fellowship and a Diversa Foundation Grant. We thank Suzanne Ratte for help with experimental technical assistance, Johanne Doleman for support with the ethical application and Daniel Gaffney for support with resources. We thank all volunteers for their participation in this study.

## Data access

Count matrices for all sequencing data DOI: 10.5281/zenodo.11105267

ArrayExpress: accession numbers for sc and bulk RNA-seq requested, assignment pending.

## Declaration/Conflict of interest

- The cups (LadyCup) for the endometriosis cohort were donated by Ladyplanet GmbH.
- DA, PCS and MB conducted this work while employed at EPFL in Switzerland. DA is currently employed by and holds Alithea Genomics company stock. MB is currentl employed by Lonza AG. PCS is currently employed by and holds F. Hoffmann-La Roche Ltd. company stock.

## Notes

### Funding Statement

This study was funded by:
- an "Early Detection Gynaecological Cancers Pump Priming award 2017" by CRUK to AG.
- HFSP and Diversa Foundation Grant to PCS.

### Author Declarations

South West - Frenchay Research Ethics Committee (IRAS project ID 221351) gave ethical approval for the healthy cohort work. Swiss Cantonal Authorities Vaud (CER-VD, project nr. 2016-00770) gave ethical approval for the endometriosis cohort work.

